# SwissPedData: Standardising hospital records for the benefit of paediatric research

**DOI:** 10.1101/2021.06.16.21258848

**Authors:** Manon Jaboyedoff, Milenko Rakic, Sara Bachmann, Christoph Berger, Manuel Diezi, Oliver Fuchs, Urs Frey, Alain Gervaix, Amalia Stefani Glücksberg, Michael Grotzer, Ulrich Heininger, Christian R. Kahlert, Daniela Kaiser, Matthias V. Kopp, Roger Lauener, Thomas J. Neuhaus, Paolo Paioni, Klara Posfay-Barbe, Gian Paolo Ramelli, Umberto Simeoni, Giacomo Simonetti, Christiane Sokollik, Ben D. Spycher, Claudia E. Kuehni

## Abstract

**Background:** Improvement of paediatric healthcare is hampered by inefficient processes of generating new evidence. Clinical research often requires extra encounters with patients, is costly, takes place in an artificial situation with a biased selection of patients, and entails long delays until new evidence is implemented into health care. Electronic health records (EHR) contain detailed information on real patients and cover the entirety of patients. However, the use of EHR for research is limited because they are not standardized between hospitals. This leads to disproportionate amounts of work for extracting data of interest and frequently data are incomplete and of poor quality.

**Aims:** SwissPedData aims to lay the foundation for a paediatric learning health system in Switzerland by facilitating EHR-based research. In this project, we aimed to assess the way routine clinical data are currently recorded in large paediatric clinics in Switzerland and to develop a national EHR-based common data model (CDM) that covers all processes of routine paediatric care in hospitals.

**Methods:** A taskforce of paediatricians from large Swiss children’s hospitals reviewed the current status of routine data documentation in paediatric clinical care and the extent of digitalization. We then used a modified Delphi method to reach a broad consensus on a national EHR-based CDM.

**Results:** All Swiss children’s hospitals use EHR to document some or all aspects of care. 119 paediatricians, representing eight hospitals and all paediatric subspecialties, participated in an extended Delphi process to create SwissPedData. The group agreed on a national CDM that comprises a main module with general paediatric data and sub-modules relevant to paediatric subspecialties. The data dictionary includes 336 common data elements (CDEs): 76 in the main module on general paediatrics and between 11 and 59 CDEs per subspecialty module. Among these, 266 were classified as mandatory, 52 as recommended and 18 as optional.

**Conclusion:** SwissPedData is a CDM for information to be collected in EHR of Swiss children’s hospitals. It covers all care processes including clinical and paraclinical assessment, diagnosis, treatment, disposition and care site. All participating hospitals agreed to implement SwissPedData in their clinical routine and clinic information systems. This will pave the way for a national paediatric learning health system in Switzerland that enables fast and efficient answers to urgent clinical questions by facilitating high-quality nationwide retrospective and prospective observational studies and recruitment of patients for nested prospective studies and clinical trials.

## Introduction

The generation of new evidence in medicine and improvement in patient care are hampered by inefficient and laborious processes [1, 2]. Most evidence is gathered through stand-alone research projects that are costly and time-consuming, and are conducted in an artificial research setting with a selected sample of patients. It also takes long until evidence is implemented in health care [3]. Delays of many years are common, caused by the need to acquire research grants, recruit staff, obtain ethical approval, set up the study, recruit participants, collect and analyse data, write up and publish results and integrate them in current standards of care. Research in the paediatric populations faces particular challenges because of small sample sizes and the fact that children represent a vulnerable patient group. Paediatric research lags behind adult research for various reasons including the facts that the paediatric population is small, many paediatric health conditions are rare and ethical requirements are high. Given these constraints, results from studies in adults are often extrapolated to children [4] [5]. However, because of the important changes that occur during their development, children differ fundamentally from adults in many aspects. These include large age-related differences in susceptibility to environmental influences, in disease manifestations, in the adequacy and performance of diagnostic tests, in drug disposition and in response to treatment [6].

The digitalization of health records could significantly improve the evidence for paediatric medicine and rare diseases as it potentially allows easy and fast access to clinical data from routine patient encounters. It could make clinical research faster, cheaper and results more representative for patients typically seen in healthcare. Electronic health records (EHR) are widely used in hospitals to document clinical and administrative information about patient’s encounters. Unfortunately, they are rarely standardized within and between institutions and data are often entered in an open text fields resulting in unstructured data. Research on rare diseases relies on data from multiple centres and is limited by the time and costs of extracting and recoding these data into a common format. Such data abstraction is particularly challenging when the original data are unstructured [7, 8]. Natural language processing and machine learning methods are increasingly being used to process unstructured data and make them available to research, however, many challenges remain [9]. Furthermore, retrospective standardisation often leads to a loss of information and impairment of data quality. These limitations could largely be circumvented if the original data were recorded in a structured and standardized way [10, 11]. A common architecture of EHR allowing structured data capture during routine medical encounters could allow rapid analytics of healthcare data followed by speedy feedback of generated knowledge into the same health care settings, a process called a learning health system [12, 13].

The Swiss Personalized Health Network (SPHN), an initiative of the Swiss Federal Government, aims to achieve a nationwide interoperability of health-data produced in university hospitals (https://sphn.ch). SPHN funds the development of infrastructures that make health-data shareable for research, following a decentralized approach where data remain in each hospital. Data sharing should become possible either through the direct transfer of individual health-data or through distributed analyses, whereby data do not travel, but are decentrally processed by algorithms and only data summaries and results are transferred to a central location [14]. SwissPedNet, the research network of Swiss Children’s hospitals (https://www.swisspednet.ch/home/) received an infrastructure grant from SPHN to develop a common data structure in paediatric hospitals. The underlying goal of the project, which we have named SwissPedData, was to facilitate paediatric clinical research by improving and standardizing the quality of data generated bypaediatric healthcare in Switzerland. To achieve this, we first assessed the status quo i.e. the relevant aspects of paediatric care for which data are being collected, the way these data are being recorded, and the data management systems used in the participating paediatric hospitals in Switzerland. Second, we developed and approved a standardized paediatric common data model (CDM) for EHR across Switzerland conducting a multi-stage consensus-finding process among general paediatricians and paediatric subspecialists of university and cantonal children’s hospitals. This paper describes the status quo of the project, the process of standardization and the resulting CDM: SwissPedData, Version 1.0.

## Methods

### SwissPedData taskforce

SwissPedData was launched as an initiative of SwissPedNet, the clinical research network of Swiss Children’s hospitals and is supported by the Swiss Society of Paediatrics (https://www.paediatrieschweiz.ch). It is coordinated by a taskforce that consists of a core team at the Institute of Social and Preventive Medicine, University of Bern (ISPM Bern), and representatives from all participating hospitals (Figure 1). All university hospitals (Basel, Bern, Geneva, Lausanne, and Zurich) and three cantonal children’s hospitals (Lucerne, St. Gallen, and Ticino) participated. The clinic directors of each hospital proposed one senior physician to represent the management board of the hospital, and one junior physician to represent the house officers and registrars who enter most data into the EHR. Directors also suggested senior physicians representing general paediatrics and all major paediatric subspecialties for collaboration in the Delphi panel. Distinct panels for the following subspecialties were set up: paediatric cardiology, endocrinology, gastroenterology, allergy/immunology, infectious diseases, metabolic diseases, nephrology, neurology, pulmonology and rheumatology. Paediatric oncology and neonatology were considered separately, because standardized datasets for these subspecialties have already been developed by the Swiss Neonatal Network & Follow-Up Group (SwissNeoNet, https://www.neonet.ch/swissneonet) [15] and the Childhood Cancer Registry (https://www.childhoodcancerregistry.ch) [16]. Both datasets have been in use for many years and have been continuously refined and thus could be included directly in SwissPedData without further discussions. A related project is developing a CDM for Paediatric Emergency Medicine using the same approach. The results of that effort will be reported separately.

**Figure 1:**
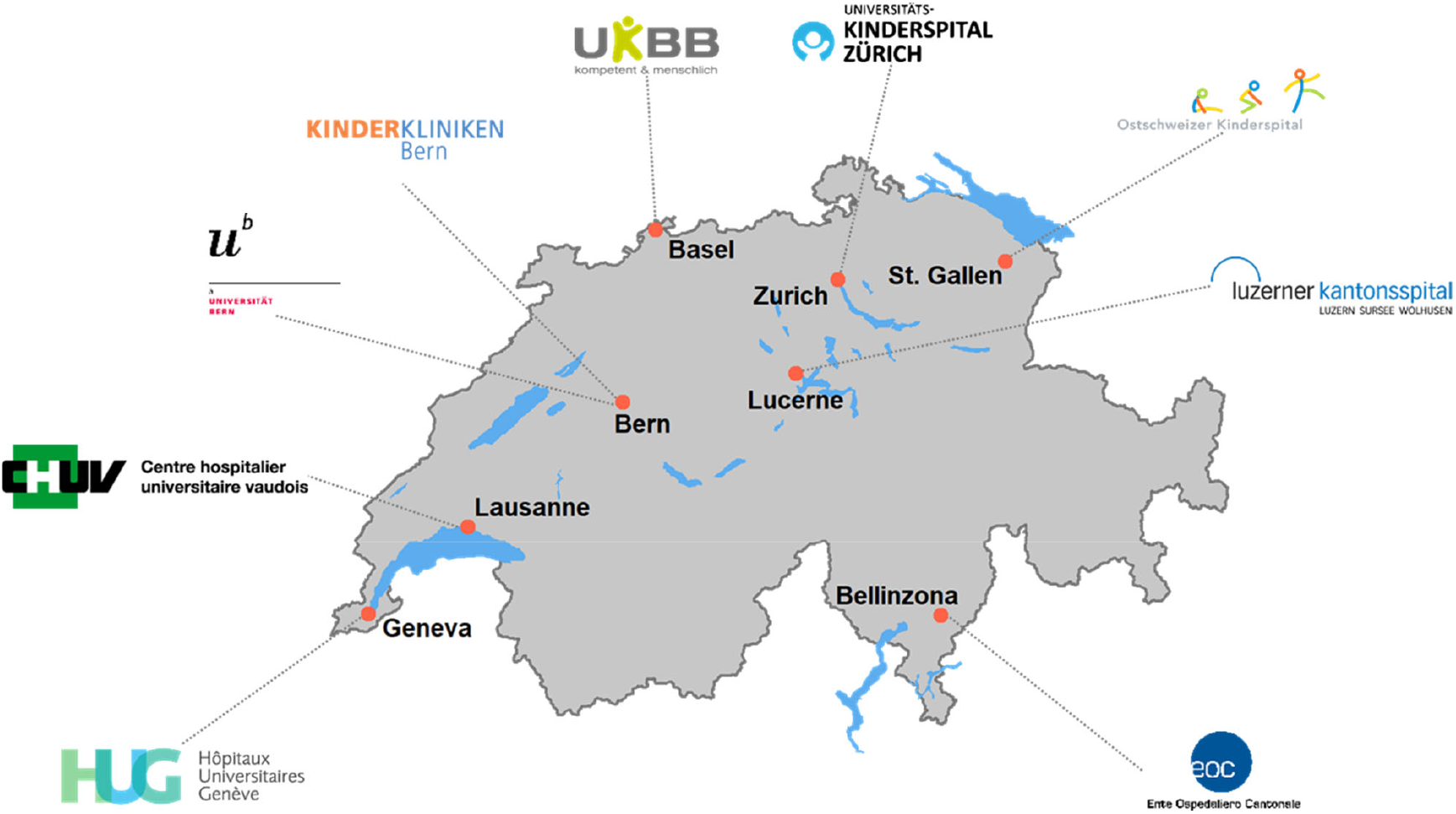
SwissPedData taskforce

### SwissPedData scope

SwissPedData focuses on the standardization of the documentation of clinical encounters by paediatricians in children’s hospitals, which encompasses medical history, physical examination, investigations, diagnosis, treatment and procedures. It excludes laboratory data and biospecimens, as this type of data is usually not entered in EHR by the clinicians themselves. Other SPHN-funded projects are working toward harmonization of laboratory data in Switzerland (https://sphn.ch/fr/network/project-overview/).

### Preparatory steps

To prepare the ground for the new CDM, the core team assessed the current status of clinical data documentation during routine encounters in participating hospitals and in ongoing clinical registries and cohort studies, and searched the literature for other initiatives aiming to standardize paediatric EHR (Figure 2). The core team visited each participating hospital and collected clinical data entry forms and information on the EHR system used and on the degree of digitalization of health records. The team identified any large existing national or regional clinical paediatric registries and cohort studies via the registry centre (https://www.paediatrieschweiz.ch/swisspedregistry/) and the clinical hubs of SwissPedNet, and through information obtained from the task force members of participating hospitals. The core team collected metadata describing the datasets collected in these registries and cohort studies and investigated the content and format of the variables.

**Figure 2:**
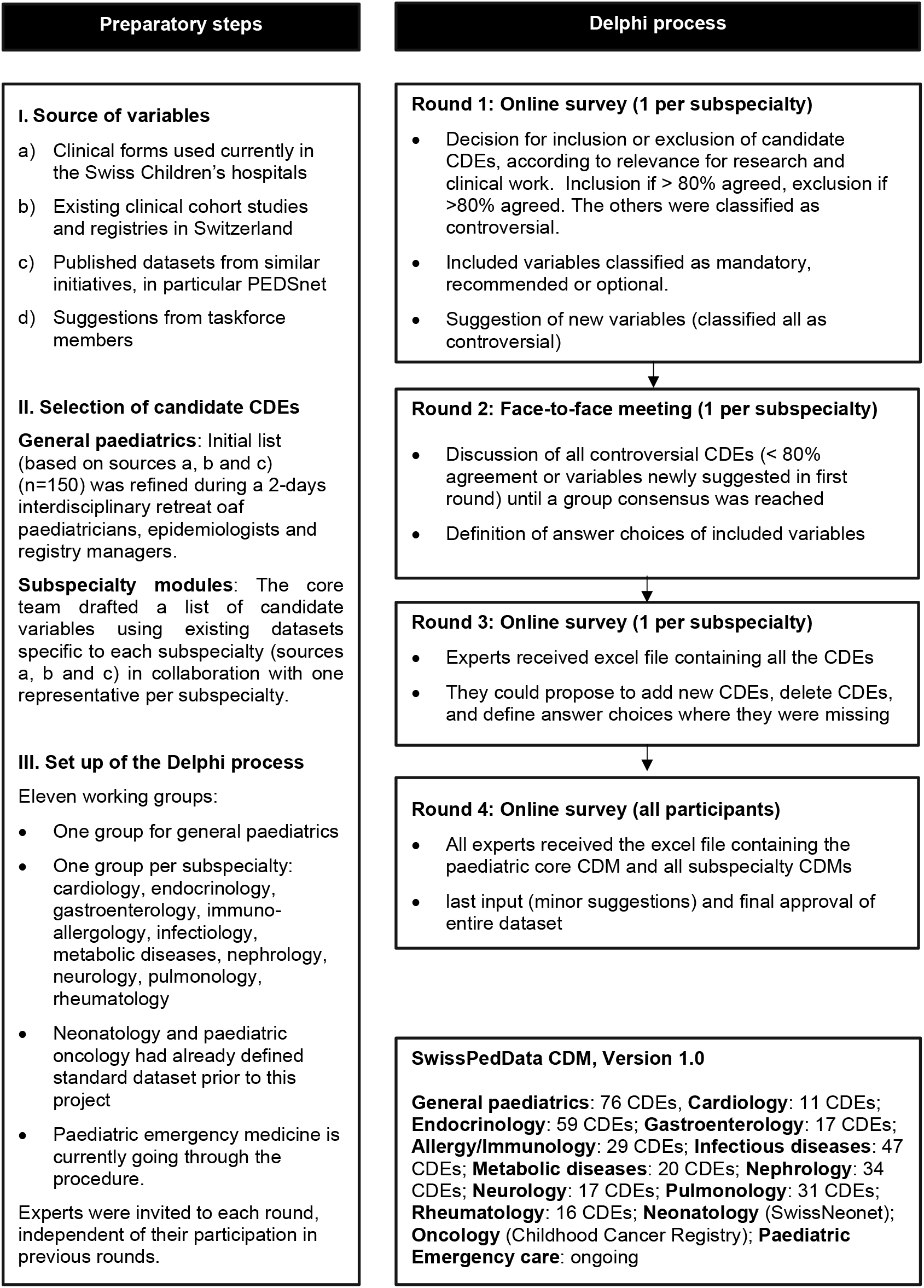
Consensus finding process followed to define SwissPedData, a common data model for recording routine encounters in children’s clinics in Switzerland. CDE: Common Data Element.

The core-team also conducted a non-systematic, focused literature search to identify approaches to standardize paediatric data across multiple centres in other countries. Reference list of the relevant publications identified were also scanned.

### Selection of candidate common data elements for SwissPedData

Based on the information gained in the preparatory phase, the core team defined an initial list of common data elements (CDEs) to be considered for inclusion in the main module (general paediatrics) of SwissPedData. This was done based on an overview of the clinical data routinely documented in the hospitals; the variables collected in ongoing clinical cohort studies and registries; and the datasets of similar international initiatives. The initial list of CDEs was further refined during a 2-days retreat held at the ISPM Bern in an interdisciplinary group including six paediatricians, three paediatric epidemiologists and two paediatric registry managers.

For paediatric subspecialties, the initial list of candidate CDEs was drafted by the core team together with one hospital paediatrician who represented the subspecialty. This first draft was based on existing datasets specific to each subspecialty, such as large cohort studies or clinical registries, and/or on expert opinion (Figure 2, Selection of candidate CDEs).

### Reaching a consensus: the Delphi process

The consensus finding process aimed to reach agreement on 1) a list of CDEs for SwissPedData, 2) standardized answer format for each CDE and 3) a classification of each CDE as either mandatory, recommended or optional. Starting with the initial selection of candidate CDEs, we implemented four Delphi rounds consisting of one face-to-face meeting and three online surveys to obtain a final CDM based on a wide consensus (Figure 2). The Delphi method achieves consensus in a multi-round iterative process that involves eliciting opinions from experts and controlled feedback from the coordinating team [17, 18]. The same basic scheme was followed for the main general paediatric module and each of the subspecialty modules. All experts were invited again to each round, irrespective of whether or not they had given their input in the previous rounds. For each online survey, experts were requested to complete the questionnaire within two weeks. Those who had not responded within one week received a reminder e-mail. The online surveys were programmed with the software SurveyMonkey Inc., San Mateo, California, USA and analysed using Microsoft Excel.

**In the first round**, experts evaluated the candidate CDEs according to their relevance for research and clinical work (Figure 2, Round 1). Each expert was asked to vote for inclusion or exclusion of each candidate CDE and to suggest any additional CDEs. When opting for inclusion of a CDE, experts were further asked to classify the CDE as: “mandatory”, “recommended” or “optional”. We retained CDEs that reached 80% for inclusion (designated as agreed) and excluded CDEs for which 80% of experts voted for exclusion. All other CDEs, including the additional CDEs suggested by experts, were classified as “controversial”. In the literature, there is no standard level of consensus but levels ranging from 50% to 80% are commonly used [19, 20].

**The second round** consisted of face-to-face meetings which were moderated by the core team and held at the ISPM Bern. During the face-to-face meetings, participants discussed all controversial CDEs and the additional CDEs suggested in the first online survey. They also agreed on standardized answer formats for included CDEs. Eligible answer formats were a date, a date and time, a number, a binary response (e.g. yes/no), standardized response options or free text. When the discussions did not lead to a consensus, we used majority voting. Each face-to-face meeting lasted about three hours.

**The third round** was again an e-Delphi survey, with participants being asked to check if key CDEs were missing in their discipline, and to propose standardised answer formats or response options where these were missing.

In the **fourth and final round**, the agreed CDMs and answer formats were sent by email to all experts for last inputs and final approval.

Ethical approval was not required for this study which did not involve the collection or use of patient’s data.

## Results

### Current status of EHR in participating hospitals and existing initiatives aiming to standardize paediatric data

The eight participating hospitals were using different clinical systems for EHRs from various vendors (Table 1). Their degree of digitalisation varied: While some hospitals were using EHR for all processes of care, others were doing so only for some. For example, all hospitals were recording clinical notes relating to inpatients electronically but only half of the hospitals were using electronic drug prescriptions at the time of the survey.

**Table 1:**
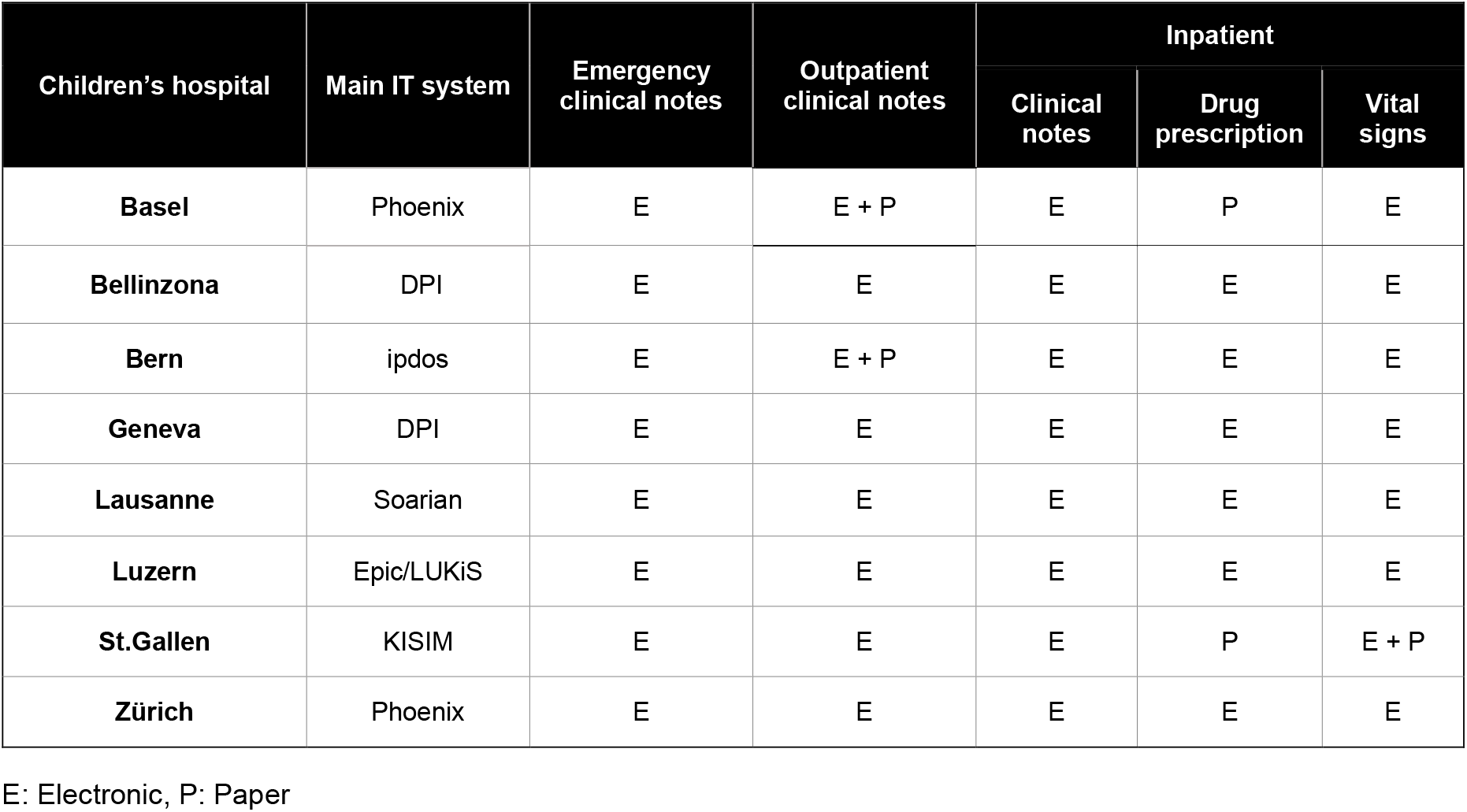
Overview of Electronic Health Records Systems in Swiss children’s hospitals, and degree of digitalization of clinical documentation

| Children's hospital | Main IT system | Emergency clinical notes | Outpatient clinical notes | Inpatient |  |  |
| --- | --- | --- | --- | --- | --- | --- |
|  |  |  |  | Clinical notes | Drug prescription | Vital signs |
| **Basel** | Phoenix | E | E + P | E | P | E |
| **Bellinzona** | DPI | E | E | E | E | E |
| **Bern** | ipdos | E | E + P | E | E | E |
| **Geneva** | DPI | E | E | E | E | E |
| **Lausanne** | Soarian | E | E | E | E | E |
| **Luzern** | Epic/LUKiS | E | E | E | E | E |
| **St.Gallen** | KISIM | E | E | E | P | E + P |
| **Zürich** | Phoenix | E | E | E | E | E |

E: Electronic, P: Paper

We identified 5 paediatric cohort studies and 25 paediatric clinical registries with a nationwide or multiregional reach (Appendix 1). The focused literature search identified four projects with similar goals in other countries, namely PECARN (Pediatric Emergency Care Applied Research Network), PHIS+ (Pediatric Health Information System), PROS (Pediatric Research in Office Settings) and PEDSnet. The initiative most similar to ours was PEDSnet, an American national Paediatric learning health system that was founded in 2014 by eight Children’s Hospitals primarily to obtain child-specific data on efficacy and safety of new and approved drugs [21] (https://pedsnet.org/data/). Currently, PEDSnet hosts analysis-ready standardised longitudinal data from primary, secondary and tertiary care of over 6.5 million patients. PEDSnet uses a common interoperable data platform that optimizes the use of EHR ensuring that data are entered once only. The collected data includes demographics, vital status, encounters, diagnoses, vital signs, treatment, immunizations, among others (https://pedsnet.org/data/common-data-model/).

### Consensus finding process (Delphi method)

Clinic directors proposed 121 experienced general paediatricians and subspecialists for the Delphi process of whom 119 agreed to participate. Of these, 73 took part in the first round (online survey), 45 attended the second round (face-to-face meetings), 58 commented in the third round of the Delphi process and 68 gave their final approval for the dataset (Appendix 2). Working groups included between 7 and 14 members. All disagreements could be settled during the process through majority voting or through discussions. Most disagreements were about answer format rather than about which CDEs should be included in SwissPedData.

### SwissPedData (Version 1.0): approved CDM

The resulting common data model (CDM) of SwissPedData consists of 336 CDEs: 76 in the main module on general paediatrics, and between 11 and 59 in each of the 10 subspecialty modules (Table 2 and Appendix 3). The main module covers aspects concerning all paediatric patients, whether they are outpatients or inpatients. Subspecialty modules cover aspects specific to paediatric subspecialties that are not already covered by the main module. Each module is formally structured into the same 9 domains representing all processes of care: 1. Care site, 2. Demographics, 3. Medical history, 4. Physical examination, 5. Clinical scores, 6. Investigations, 7. Diagnosis, 8. Treatment, 9. Equipment and procedures. These represent domains commonly covered by EHRs. The Care Site domain contains administrative data related to the hospital and to patient encounters. It includes type of admission, length of stay, scheduled follow-up. The Demographics domain contains demographic data, for example date of birth, gender, address, country of birth. The Medical History and the Physical Examination domains include clinical information such as birth history, family history, symptoms, medications, vital signs. The Clinical Scores domain contains specific scores for example triage scale for emergency department patients or developmental tests. The Investigations domain contains data on investigations performed, such as lung functions, renal ultrasound or blood glucose monitoring for patients with diabetes. The Diagnosis domain include diagnosis and date of diagnosis as well as diagnosis classification such as Online Mendelian Inheritance in Man (OMIM) codes. The Treatment domain contains data on medications prescribed and administered in hospital, treatment adverse events, and reason for discountinuation of treatment. The Equipment and Procedures domain countains data on procedures performed on the patient, such as dialysis.

**Table 2:**
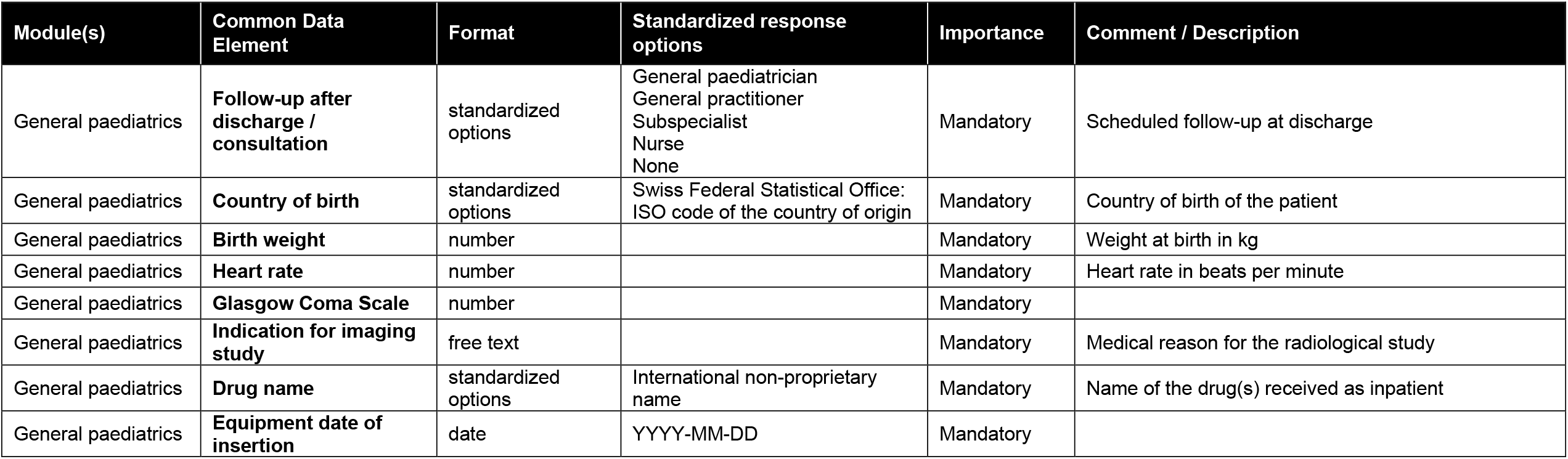
Examples of common data elements (CDE) of the core-module (general paediatrics) of SwissPedData’s common data model (CDM)

The full CDM is shown in the Appendix 3. It provides a complete list of all agreed CDE’s, their description, answer format and standardized response options, and importance (mandatory; recommended or optional). Answer choices are number, binary or standardized options, or free text. When the format “standardized option” is used, specific value sets are defined. CDEs will be implemented in children’s hospital EHR depending on their importance, categorized as: mandatory; recommended or optional. Mandatory CDEs must be implemented in EHR by all participating hospitals. Recommended CDEs should be implemented and optional CDEs may be implemented at the discretion of each hospital.

Examples of mandatory CDEs are vital parameters in the main module (general paediatrics) or “route of feeding” in the gastroenterology module. In the latter case, “route of feeding” will be recorded with standardized response options (Oral, Gastrostomy, Naso/orogastric tube, Intravenous, Other). An example for a recommended CDE is “Seizure type according to the ILEA 2017 classification of seizures” in the neurology module. “Opening Pressure at Lumbar Puncture” is an optional CDE in the same module (Appendix 3).

## Discussion

We developed SwissPedData, a standardized national CDM designed to collect clinical data during paediatric routine encounters in a harmonized way. It is the result of a broad consensus between general paediatricians and paediatric subspecialists from eight university and cantonal children’s hospitals in Switzerland. It describes all processes of paediatric medical care including clinical and paraclinical assessment, diagnosis, treatment, disposition and care site. Each part of the dataset follows the usual structure of the EHR to allow easy implementation.

### Clinical data standardization for a Swiss Paediatric learning health care system

SwissPedData aimed to standardize items up-front at the point of data entry. A prospected standardized recording of routine clinical encounters avoids duplicate entry into research databases. However, this should not happen at the expense of an increase in documentation time by clinicians, a concern raised during our Delphi process. To avoid this pitfall, we focused primarily on data elements that are not only useful for research but also for clinical work and included CDEs that are routinely documented in paediatric EHRs. SwissPedData is not comprehensive and much of the clinical documentation will remain unstandardized to preserve the rich narrative details that are difficult to capture in standardised fields but are nevertheless important for daily clinical work. These narrative data could be used by researchers applying text-mining approaches. SwissPedData could also be supplemented by questionnaires to patients and their families. The implementation of SwissPedData in EHR will include careful attention to clinician workflow to minimize potential negative consequences of standardization.

SwissPedData is designed to provide a basis for a paediatric learning health system in Switzerland, in which clinical data from different children’s hospitals can be combined to rapidly generate new knowledge relevant for day-to-day practice and translate it into improved health care for children. Existing learning health systems in other countries, such as PEDSnet in the US, have demonstrated that a paediatric learning health system can improve health outcomes of children [22, 23]. Examples include the rapid identification of children suffering from glomerular diseases for clinical trials [24], comparing weight loss and safety among bariatric procedures using EHR data [25], and recently describing the epidemiology of pediatric patients infected by SARS-CoV-2 [26].

### Strengths and limitations

The main strength of SwissPedData is that it is based on a wide agreement between pediatricians from all university and cantonal paediatric clinics in Switzerland. The project received strong support from all clinic directors of Swiss children’s hospitals, from the paediatric research Network SwissPedNet and from more than 100 experienced paediatricians who participated in its development. SwissPedData emphasizes the prospective collection of standardized data, which can greatly reduce the time and costs needed for data preparation and analysis as it avoids the need for retrospective standardisation or double entry. Our consensus finding approach can be adapted for use other medical specialties that wish to define CDM in the future.

SwissPedData has a number of omissions that are intentional. We focused on standardising a minimal set of items that are particularly relevant and specific to paediatric routine care. SwissPedData does not intend to replace existing terminologies for clinical health care such as SNOMED-CT or LOINC. Second, SwissPedData does not include laboratory data nor detailed radiologic data. However, other projects in the SPHN are working on the standardization of these domains. Third, it will need to be translated into the Swiss national languages before implementation in children’s hospitals EHR.

### SwissPedData is adapted to the Swiss context

The Swiss Health system is decentrally structured, with cantons being responsible for the organisation of local healthcare, and therefore is highly heterogenous. As a consequence, children’s clinics are relatively small with a catchment area of a few 100,000 children. Obtaining sufficient patients samples for research is only possible by combining data from multiple hospitals, particularly for rare conditions. However, given the differences in EHRs and IT systems between hospitals, this results in long delays and huge costs for obtaining, extracting, standardizing and cleaning the heterogeneous data. SwissPedData, once implemented in all children’s clinics, will allow researchers to identify and recruit patients for clinical trials in real-time, conduct retrospective studies with high-quality data and conduct nested prospective studies.

### Comparison with other projects

SwissPedData is closely aligned with PEDSnet, a US-based paediatric clinical data research network [21]. PEDSnet includes eight children’s hospitals that provide care for 2.8% of the paediatric population in the USA (2.1 million patients) [21]. The database contains standardized clinical data from electronic health records (EHR) covering 6.5 million children (https://pedsnet.org/) which forms the basis of a high-quality research program and learning health system. Studies based on PEDSnet data cover a wide range of research topics and study designs in paediatrics including descriptive epidemiology [27], computable phenotyping [24], longitudinal observational studies [28], and comparative effectiveness [29].

PEDSnet may also serve as a role model for implementation of SwissPedData and has already demonstrated its usefulness for observational and interventional research, and standardizing of care processes. Each hospital that participates in PEDSnet regularly extracts the standardized data from its EHR in a predefined way [21].

Another notable example of harmonized clinical datasets in pediatrics is the Pediatric Emergency Care Applied Research Network (PECARN), an EHR-based registry that has harmonized data in the paediatric emergency setting in 7 american paediatric emergency departments to make it usable for paediatric research. PECARN uses data resources from seven paediatric emergency departments of four hospitals [30].

### Outlook and next steps

All participating hospitals are committed to implement SwissPedData in their EHRs by 2024. A committee of clinicians and IT specialists in each hospital will supervise the implementation process. Practically, it means that EHR as seen by the users (physicians) will include the CDEs of SwissPedData. SwissPedData is intended to be evolutive and adaptive to existing needs. The CDM can be expanded to cover more domains or to include more CDEs per domain. Temporary CDEs can be added for nested research projects. Self-completed or parent-completed questionnaires can add information relating to the childs family and home environment, which is not routinely recorded in EHR. Data from primary care encounters could in future also be integrated.

In ongoing work, other prerequisites for implementation of SwissPedData are being put into place: a general consent form for use of the data from patients and caregivers, a data transfer and use agreement (DTUA) between the clinics, and protocols for obtaining ethics approval for SwissPedData overall, and for individual research projects. Some aspects are being dealt with in other infrastructure development projects of the SPHN network (www.sphn.ch), namely the C3-Study (citizen centered consent) project and the E-General Consent project. Furthermore, the SPHN provides legal agreement templates including DTUA and an ethical framework for all its projects. It is important to stress that only data useful for the clinical management of the patient will be recorded and that these data will always be stored by each children’s hospital as part of the patient’s file. The only difference to the previous procedure is that some of these clinical data will be recorded in a standardized way. To have access to these data for research, researchers will have to get an ethical approval as usual.

It is planned that SwissPedData can be implemented as a project on the SPHN infrastructure for data exchange, so that data can in future be accessed through a central portal. The SPHN Data Coordination Centre and BioMedIT (https://sphn.ch/network/projects/biomedit/) can provide assistance and infrastructure for this. An additional central coordination centre for paediatric research should facilitate communication between children’s clinics, international research partners and funders, and assist researchers in writing grant applications, obtaining ethical approval and the necessary datasets. Resources needed to maintain SwissPedData will require support of a central coordination center which encompasses an experienced researcher with ideally a background in paediatrics, an IT specialist, and local support of responsible clinicians and IT specialists in each hospital. Funding for implementation and maintenance of SwissPedData will need to be secured. Potential funding sources are participation in suitable calls for proposals, charging cost-covering fees for services provided by SwissPedData and collaboration with industry, for example for post-marketing studies. Collaboration with international partners, such as PEDSnet, are forseen and first exchanges have occurred.

In conclusion, SwissPedData defines a common data model (CDM) for clinical paediatric care based on a wide agreement among university and cantonal pediatric hospitals in Switzerland. With SwissPedData, Swiss children’s hospitals will be able to provide researchers standardized, high quality routine clinical paediatric data in a near future. SwissPedData will provide the basis for a learning health system for paediatric care in Switzerland.

## Data Availability

The SwissPedData Common Data Model is available as appendix

## Acknowledgments

We thank all the experts who participated in the Delphi process, SwissPedNet, College A, the Swiss Personalized Health NetworkSPHN, ISPM Bern staff: Alexander Laemmle, Alexander Moeller, Alexandra Wilhelm-Bals, Alexandre Datta, Alice Koehli, Andrea Duppenthaler, Andreas Nydegger, Andreas Worner, Anita Rauch, Anna Wefers, Anne Tscherter, Arnaud Merglen, Barbara Goeggel Simonetti, Juerg Barben, Birgit Donner, Caroline Roduit, Christian Braegger, Christian Kahlert, Christian Korff, Christian Huemer, Christian Lovis, Christina Schindera, Christoph Aebi, Christoph Berger, Christophe Folly, Christoph Rudin, Christian Balmer, Cristina Ardura, Claudia Boettcher, Constance Barazzone-Argiroffo, Corinna Leoni Foglia, Dagmar L’Allemand, Daniel Konrad, Daniel Trachsel, Daniela Marx-Berger, Diana Ballhausen, Dirk Fischer, Dominik Stambach, Eliane Roulet, Elvira Cannizzaro, Emanuela Valsangiacomo, Eva Pedersen, Federica Vanoni, Felicitas Bellutti, Florian Bauder, Florence Barbey, Florian Singer, François Cachat, Franziska Kunz, Gabor Szinnai, Georg Marx, Giovanni Ferrari, Gianluca Gualco, Guido Laube-Bless, Hans Peter Kuen, Hassib Chehade, Ilse Kern, Isabel Bolt, Isabelle Rochat, Jana Pachlopnik Schmid, Jean-Baptiste Armengaud, Jean-Christoph Caubet, Joan Carles Suris Granell, Joël Fluss, Johannes Spalinger, Julien Caccia, Jürg Hammer, Kanetee Busiah, Katrin Heldt, Katharina Flandera, Kristina Keitel, Laetitia Marie Petit, Lisa Kottanattu, Lorenzo Zgraggen, Luca Garzoni, Matthias Horn, Maria Otth, Matthias Baumgartner, Matthias Gautschi, Maura Zanolari-Calderari, Maurice Beghetti, Melanie Hess, Michael Hauschild, Michael Buettcher, Michael Hofer, Mirjam Dirlewanger, Myrofora Goutaki, Nicolas Regamey, Nicolas Waespe, Nicole Sekarski, Nicole Ritz, Noémie Wagner, Oliver Niesse, Oswald Hasselmann, Paloma Parvex, Paolo Tonella, Paolo Paioni, Pascale Wenger, Peter Weber, Philip Broser, Philipp Agyman, Philip Do Canto, Philippe Steenhout, Philippe Eigenmann, Piero Balice, Pierre-Alex Crisinel, Raoul Furlano, Rebeca Mozun, Regula Laux, Regina Wespi, Robert Steinfeld, Sabine Pallivathukal, Sandra Asner, Sebastian Grunt, Sébastien Lebon, Sébastien Papis, Selina Pinosch, Sibylle Tschumi, Stefano di Bernardo, Sylvain Blanchon, Thomas Schmitt-Mechelke, Ulrike Halbsguth, Urs Zumsteg, Valérie Schwitzgebel, Valérie McLin, Verena Pfeiffer, Yacine Aggoun

## Address for correspondence including e-mail address

Prof. Dr. med. Claudia Kuehni, Institute of Social and Preventive Medicine, University of Bern, Bern, Switzerland.

## Appendixes

**Appendix 1:** Paediatric registries and cohort studies in Switzerland

**Appendix 2:** Number of experts involved at each stage of the Delphi process

**Appendix 3:** SwissPedData CDM version 1.0

## List of abbreviations

CDE: Common data element
CDM: Common data model
EHR: Electronic Health record
ISPM Bern: Institute of Social and Preventive Medicine, University of Bern
PEDSnet: A multi-specialty network that conducts observational research and clinical trials across multiple children’s hospital health systems in the US (www.pedsnet.org)
PECARN: Pediatric Emergency Care Applied Research Network
SPHN: Swiss Personalized Health Network (https://sphn.ch/)
SwissPedData: “Harmonising the collection of health-related data and biospecimens in paediatric hospitals throughout Switzerland”, an infrastructure development project of the SPHN funded in 2017
SwissPedNet: Swiss Research Network of clinical Pediatric Hubs (www.swisspednet.ch)

