## Appendix 1 for "SwissPedData: Standardising hospital records for the benefit of paediatric research"

**Appendix 1:** Paediatric registries and cohort studies in Switzerland

| Registry / Cohort Study | Coverage |
|---|---|
| Childhood Cancer Registry ChCR | National |
| Swiss Primary Ciliary Dyskinesia Registry (SPCDR) | National |
| Swiss Cerebral Palsy Registry (Swiss-CP-Reg) | National |
| Swiss Growth Registry (SGR) | National |
| Swiss Paediatric Airway Cohort (SPAC) | National |
| Swiss Paediatric Renal Registry (SPRR) | National |
| Swiss Rare Disease Registry (SRDR) | National |
| Swiss Registry for Neuro-Muscular Disorders (Swiss-Reg-NMD) | National |
| Cystic Fibrosis (CF) newborn screening | National |
| Juvenile Inflammatory Rhumatism cohort (JIRcohorte) | European |
| SwissNeoNet Minimal Neonatal Data Set (MNDS) | National |
| SwissNeoNet National Asphyxia and Cooling Registry (ASP) | National |
| SwissNeoNet Follow-Up (FU) | National |
| Swiss NeuroPaediatric Stroke Registry (SNPSR) | National |
| Swiss Congenital Lung Anomalies (CLA) Registry | National |
| Swiss Mother and Child HIV Cohort Study (MoCHiV) | National |
| Swiss Cystic Fibrosis Infant Lung Development (SCILD) cohort | National |
| Swiss Pediatric Surveillance Unit (SPSU) | National |
| Swiss Hemophilia Registry (SHN) | National |
| COST Action BM1105 Patient Registry - GnRH Network | European |
| Registry of congenital anomalies in the canton of Vaud | National |
| Swiss Cleft lip and Palate Registry | National |
| Swiss Biliary Atresia Registry | National |
| Swiss registry on Autoimmune Hepatitis | National |
| European Registry for Primary Immunodeficiencies (ESID registry) | European |
| European Cystic Fibrosis Patient Registry (ECFSPR) | European |
| European Childhood Interstitial Lung Disease (chILD-EU) Registry | European |
| Swiss Inflammatory Bowel Disease Pediatric Cohort Study (Swiss IBD Pediatric Cohort Study) | National |
| Splenectomy Registry | Global |
| Pediatric and Adult Intercontinental Registry on Chronic ITP (PARC-ITP registry) | Global |
| Diabetes Patienten Verlaufsdokumentation Registry (DPV) | European |
