## Appendix 2 for "SwissPedData: Standardising hospital records for the benefit of paediatric research"

**Appendix 2:** Number of experts involved at each stage of the Delphi process

| Paediatric specialty | Number of experts invited | Delphi process, number of experts involved | | | |
|---|---|---|---|---|---|
| | | 1st round | 2nd round | 3rd round | 4th round |
| **General paediatrics** | 14 | 8 | 4 | 4 | 5 |
| **Cardiology** | 13 | 10 | 4 | 7 | 8 |
| **Endocrinology** | 12 | 7 | 6 | 8 | 9 |
| **Gastroenterology** | 10 | 8 | 4 | 4 | 6 |
| **Allergy/Immunology** | 12 | 6 | 4 | 8 | 7 |
| **Infectiology** | 11 | 8 | 5 | 6 | 9 |
| **Metabolic diseases** | 8 | 7 | 2 | 4 | 3 |
| **Nephrology** | 12 | 3 | 4 | 5 | 4 |
| **Neurology** | 14 | 5 | 4 | 3 | 5 |
| **Pulmonology** | 11 | 8 | 5 | 4 | 6 |
| **Rheumatology** | 8 | 3 | 3 | 5 | 6 |
