## Appendix 3 for "SwissPedData: Standardising hospital records for the benefit of paediatric research"

**Appendix 3** : SwissPedData Common Data Model (CDM), Version 1.0

| Module(s) | Common Data Element | Format | Standardized response options | Importance | Comment / Description |
|---|---|---|---|---|---|
| **Domain: Care site** | | | | | |
| General paediatrics | **Type of admission** | standardized options | Elective admission<br>Emergency admission | Mandatory | |
| General paediatrics | **Provenance** | standardized options | Other hospital<br>Emergency department<br>Home<br>Other | Mandatory | |
| General paediatrics | **Care Handling Type** | standardized options | Inpatient<br>Outpatient | Mandatory | |
| General paediatrics | **Visit start date and time** | datetime | YYYY-MM-DD hh:mm:ss | Mandatory | Datetime at which the interaction between individual and the care provider institute started |
| General paediatrics | **Visit end date and time** | datetime | YYYY-MM-DD hh:mm:ss | Mandatory | Datetime at which the interaction between individual and the care provider institute stopped |
| General paediatrics | **Datetime of admission** | datetime | YYYY-MM-DD hh:mm:ss | Mandatory | Datetime of patient's admission to the care provider institute |
| General paediatrics | **Discharge destination** | standardized options | Home<br>Other hospital<br>Institution<br>Other | Mandatory | Location to which the patient is discharged |
| General paediatrics | **Follow-up after discharge / consultation** | standardized options | General paediatrician<br>General practitioner<br>Subspecialist<br>Nurse<br>None | Mandatory | Scheduled follow-up at discharge |
| General paediatrics | **Translator needed** | standardized options | Yes<br>No<br>Unknown | Recommended | Translator needed for communication between patient and healthcare team |
| General paediatrics | **Hospital** | standardized options | See comments | Mandatory | Standardized response options will be name of participating children’s hospitals |
| General paediatrics | **Department** | standardized options | See comments | Mandatory | Standardized response options will be name of departments of participating children’s hospitals |
| General paediatrics | **Unit** | standardized options | See comments | Mandatory | Standardized response options will be name of units of participating children’s hospitals |
| Infectious diseases | **If coming from another hospital: Country** | standardized options | Swiss Federal Statistical Office: ISO code of the country of origin | Mandatory | Country of originating hospital |
| **Domain: Demographics** | | | | | |
| General paediatrics | **Patient Datetime of birth** | datetime | YYYY-MM-DD hh:mm:ss | Mandatory | Datetime of birth of the patient |
| General paediatrics | **Country of birth** | standardized options | Swiss Federal Statistical Office: ISO code of the country of origin | Mandatory | Country of birth of the patient |
| General paediatrics | **Place of birth (CH)** | number | Postal code (PLZ/NPA) | Mandatory | Municipality of birth of the patient if in Switzerland, coded by postal codes (PLZ/NPA). |

**Appendix 3** : SwissPedData Common Data Model (CDM), Version 1.0

| Module(s) | Common Data Element | Format | Standardized response options | Importance | Comment / Description |
|---|---|---|---|---|---|
| General paediatrics | **Patient administrative gender** | standardized options | Male<br>Female<br>Other | Mandatory | |
| General paediatrics | **Address (postal code)** | number | Postal code (PLZ/NPA) | Mandatory | Current address of the patient, coded by postal codes (PLZ/NPA). Exact address should also be recorded |
| General paediatrics | **Nationality** | standardized options | Swiss Federal Statistical Office: ISO code of the country of origin | Mandatory | Current nationality of the patient |
| General paediatrics | **Date of immigration** | date | YYYY-MM-DD | Mandatory | Date of first immigration to Switzerland if born abroad |
| Infectious diseases | **If immigrant: Type of residency permit** | standardized options | B<br>C<br>G<br>L<br>F<br>N<br>S<br>undocumented | Optional | |
| Metabolic diseases | **Ethnicity of the mother** | standardized options | See comments | Optional | Standard classification to be defined |
| Metabolic diseases | **Ethnicity of the father** | standardized options | See comments | Optional | Standard classification to be defined |
| Rheumatology<br>Pulmonology | **Ethnicity of the patient** | standardized options | See comments | Optional<br>Recommended | Standard classification to be defined. Optional for rheumatology, recommended for pulmonology. |
| **Domain: Medical history** | | | | | |
| General paediatrics | **Reason for consultation / for admission** | free text | | Mandatory | Main reason for consultation or for admission. Standard classification not defined. |
| General paediatrics | **Current medications: Drug name** | standardized options | International non-proprietary name | Mandatory | Name of the drug(s) received as inpatient |
| General paediatrics | **Current medications: Route of administration** | standardized options | Oral<br>Intravenous<br>Subcutaneous<br>Intramuscular<br>Intrathecal<br>Rectal<br>Inhalation<br>Cutaneous<br>Ocular<br>Nasal<br>Otic<br>Other | Mandatory | |
| General paediatrics | **Current medications: Frequency of administration** | number | | Mandatory | Number of administrations per 24 hours |
| General paediatrics | **Current medications: Dose** | number | | Mandatory | Dose given at each administration of the drug |

**Appendix 3** : SwissPedData Common Data Model (CDM), Version 1.0

| Module(s) | Common Data Element | Format | Standardized response options | Importance | Comment / Description |
|---|---|---|---|---|---|
| General paediatrics | **Current medications: Dose unit** | standardized options | | Mandatory | List of possible units to be defined |
| General paediatrics | **Use of complementary medicine** | yes/no | | Optional | Patient treated with complementary medicine at home or in hospital |
| General paediatrics | **Birth weight** | number | | Mandatory | Weight at birth in kg |
| General paediatrics | **Birth length** | number | | Mandatory | Length at birth in cm |
| General paediatrics | **Birth's head circumference** | number | | Mandatory | Head circumference at birth in cm |
| General paediatrics | **Delivery mode** | standardized options | Caesarean section<br>Instrumental vaginal delivery<br>Spontaneous vaginal delivery | Mandatory | Birth delivery mode |
| General paediatrics | **Gestational age** | number | | Mandatory | Post-menstrual age at birth in week and days |
| General paediatrics | **Apgar score 1 min** | number | | Recommended | Apgar score 1 min after birth |
| General paediatrics | **Apgar score 5 min** | number | | Recommended | Apgar score 5 min after birth |
| General paediatrics | **Apgar score 10 min** | number | | Recommended | Apgar score 10 min after birth |
| General paediatrics | **Mother's year of birth** | number | | Mandatory | Year of birth of the mother |
| General paediatrics | **Father's year of birth** | number | | Mandatory | Year of birth of the father |
| General paediatrics | **Year(s) of birth of sibling(s)** | number | | Mandatory | Year of birth of sibling(s) if any |
| General paediatrics | **Drug allergies** | standardized options | International Nonproprietary Name of drug | Mandatory | Known drug allergies |
| General paediatrics | **Documented food allergies** | yes/no | | Mandatory | Presence of any documented food allergy |
| Endocrinology | **Age at menarche** | number | | Mandatory | Age at menarche in years |
| Endocrinology | **Age at thelarche** | number | | Mandatory | Age at thelarche in years |
| Endocrinology | **Age at pubarche** | number | | Mandatory | Age at pubarche in years |
| Endocrinology | **Single/Multiple birth** | number | | Recommended | Number of children born from the same pregnancy as the patient's |
| Endocrinology | **Neonatal hypoglycaemia** | standardized options | No<br>Yes, confirmed<br>Yes, reported by patient/family | Recommended | History of hypoglycaemia in the neonatal period |
| Endocrinology | **Neonatal hyperbilirubinemia** | standardized options | No<br>Yes, confirmed<br>Yes, reported by patient/family | Recommended | History of hyperbilirubinemia in the neonatal period (only hyperbilirubinemia treated with phototherapy) |
| Endocrinology<br>Nephrology | **Mother's height** | number | | Mandatory | Height of the mother in cm |
| Endocrinology<br>Nephrology | **Father's height** | number | | Mandatory | Height of the father in cm |
| Endocrinology | **Mother's age at menarche** | number | | Mandatory | Age of the mother at menarche in years |

**Appendix 3** : SwissPedData Common Data Model (CDM), Version 1.0

| Module(s) | Common Data Element | Format | Standardized response options | Importance | Comment / Description |
|---|---|---|---|---|---|
| Endocrinology | **Father's puberty** | standardized options | Normal<br>Early<br>Late | Mandatory | |
| Endocrinology | **Diabetes in first degree relatives** | standardized options | No<br>Yes, Type 1<br>Yes, Type 2<br>Yes, Monogenic<br>Unknown | Mandatory | Any type of diabetes in a first degree relative |
| Endocrinology | **Thyroid disorder in first degree relative** | yes/no | | Mandatory | Presence of thyroid disorder in a first degree relative |
| Endocrinology | **Other auto-immune disorders in first degree relative** | yes/no | | Recommended | Presence of auto-immune disorder in a first degree relative. With added box for free text to specify the disease. |
| Endocrinology | **Other endocrinopathy in first degree relative** | yes/no | | Mandatory | Presence of endocrinopathy in a first degree relative. With added box for free text to specify the disease. |
| Endocrinology | **Fertility problems in first degree relative** | yes/no | | Recommended | Presence of fertility problems in first degree relatives. With added box for free text to specify the disease. |
| Endocrinology | **Severe hypoglycaemia (requiring assistance OR coma)** | number | | Mandatory | Number of events since last visit |
| Endocrinology | **Mild hypoglycaemia (BG < 3.9mmol/l)** | number | | Mandatory | Number of events per month |
| Endocrinology | **Ketoacidosis** | standardized options | No<br>Yes, managed ambulatorily<br>Yes, with hospitalization | Mandatory | History of ketoacidosis |
| Endocrinology | **Diagnostic of obesity in first degree relative** | yes/no | | Recommended | Diagnostic of obesity in a first degree relative |
| Gastroenterology | **Nutrition habits** | standardized options | No specific diet<br>Vegetarian<br>Vegan<br>Other | Recommended | Nutrition habits of the patient |
| Gastroenterology<br>Metabolic diseases | **Route of feeding** | standardized options | Oral<br>Gastrostomy<br>Naso/orogastric tube<br>Intravenous<br>Other | Mandatory | The route(s) by which the patient is fed |
| Allergy/Immunology | **History of rhinoconjonctivitis** | standardized options | Yes, reported<br>Yes, documented<br>No | Mandatory | |
| Allergy/Immunology | **History of atopic dermatitis** | standardized options | Yes, reported<br>Yes, documented<br>No | Mandatory | |
| Allergy/Immunology | **History of wheezing** | standardized options | Yes, reported<br>Yes, documented<br>No | Mandatory | |

**Appendix 3** : SwissPedData Common Data Model (CDM), Version 1.0

| Module(s) | Common Data Element | Format | Standardized response options | Importance | Comment / Description |
|---|---|---|---|---|---|
| Allergy/Immunology | **History of asthma** | standardized options | Yes, reported<br>Yes, documented<br>No | Mandatory | |
| Allergy/Immunology | **Respiratory support during first hours of life** | standardized options | Yes, reported<br>Yes, documented<br>No | Recommended | Presence of any kind of respiratory support (non-invasive and invasive ventilation) during first hours of life |
| Allergy/Immunology | **Supplemental O2 during first hours of life** | standardized options | Yes, reported<br>Yes, documented<br>No | Recommended | Supplemental oxygen administered during first hours of life |
| Allergy/Immunology | **Chronic diarrhea** | yes/no | | Mandatory | |
| Allergy/Immunology | **Number of hospitalisations for IV antibiotherapy in life** | number | | Mandatory | |
| Allergy/Immunology | **Maximal number of otitis media in one year** | number | | Mandatory | |
| Allergy/Immunology | **Number of pneumonias in life** | number | | Mandatory | |
| Allergy/Immunology | **Number of sinusitis in life** | number | | Mandatory | |
| Allergy/Immunology | **Number of meningitis in life** | number | | Mandatory | |
| Allergy/Immunology | **Family history of atopic diseases** | yes/no | | Mandatory | Presence of atopic diseases in a first degree relative |
| Allergy/Immunology | **Family history of immunodeficiency** | yes/no | | Mandatory | Presence of immunodeficiency in a first degree relative |
| Allergy/Immunology | **Family history of auto-immune disease** | yes/no | | Mandatory | Presence of auto-immune disease in a first degree relative |
| Allergy/Immunology | **Family history of angioedema** | yes/no | | Mandatory | Presence of angioedema in a first degree relative |
| Allergy/Immunology<br>Gastroenterology | **Documented food allergy by oral food challenge** | yes/no | | Mandatory | Presence of any documented food allergy (diagnosed by physician) |
| Allergy/Immunology | **Hymenoptera venom allergies** | standardized options | Yes, reported<br>Yes, documented<br>No | Mandatory | Known documented hymenopter allergies |
| Allergy/Immunology | **History of anaphylaxis** | standardized options | Yes, reported<br>Yes, documented<br>No | Mandatory | History of anaphylaxis |
| Allergy/Immunology | **Autoimmune or inflammatory diseases in the patient** | yes/no | | Mandatory | Classification for type of autoimmunity (organ specific or systemic) and organ(s) involved will be further defined. |
| Infectious diseases | **History of fever (>38°C)** | yes/no | | Mandatory | |
| Infectious diseases | **If history of fever: Number of days with fever** | number | | Mandatory | |
| Infectious diseases | **History of cough** | yes/no | | Mandatory | |

**Appendix 3** : SwissPedData Common Data Model (CDM), Version 1.0

| Module(s) | Common Data Element | Format | Standardized response options | Importance | Comment / Description |
|---|---|---|---|---|---|
| Infectious diseases | **History of running nose** | yes/no | | Mandatory | |
| Infectious diseases | **History of diarrhea** | yes/no | | Mandatory | |
| Infectious diseases | **History of vomiting** | yes/no | | Mandatory | |
| Infectious diseases | **History of headache** | yes/no | | Mandatory | |
| Infectious diseases | **Travel history in the last 6 months** | standardized options | Swiss Federal Statistical Office: ISO code of the country of origin (selection of >1 possible) | Mandatory | Country(ies) visited in the last 6 months |
| Infectious diseases | **History of tick bite** | yes/no | | Recommended | |
| Infectious diseases | **If history of tick bite: Month of tick bite** | date | YYYY-MM | Recommended | |
| Infectious diseases | **History of contact with animals** | yes/no | No<br>Yes | Optional | Standard animal list to be defined |
| Infectious diseases | **Pertussis immunization during pregnancy** | yes/no | | Mandatory | For patients under 6 months of age |
| Infectious diseases | **Influenza immunization during pregnancy** | yes/no | | Mandatory | For patients under 6 months of age |
| Infectious diseases | **Prolonged rupture of membranes** | yes/no | | Mandatory | For patients under 1 month of age. Prolonged rupture defined as longer than 18h |
| Infectious diseases | **Maternal GBS colonization** | standardized options | Positive<br>Negative<br>Unknown | Mandatory | For patients under 1 month of age |
| Infectious diseases | **Maternal HIV serology** | standardized options | Positive<br>Negative<br>Unknown | Mandatory | For patients under 1 month of age |
| Infectious diseases | **Maternal HBsAg** | standardized options | Positive<br>Negative<br>Unknown | Mandatory | For patients under 1 month of age |
| Infectious diseases | **Maternal HBsAb** | standardized options | Positive<br>Negative<br>Unknown | Mandatory | For patients under 1 month of age |
| Infectious diseases | **Maternal HBcAb** | standardized options | Positive<br>Negative<br>Unknown | Mandatory | For patients under 1 month of age |
| Infectious diseases | **Maternal HBeAg** | standardized options | Positive<br>Negative<br>Unknown | Mandatory | For patients under 1 month of age |
| Infectious diseases | **Maternal HCV serology** | standardized options | Positive<br>Negative<br>Unknown | Mandatory | For patients under 1 month of age |
| Infectious diseases | **Maternal CMV serology (IgG / IgM)** | standardized options | Positive<br>Negative<br>Unknown | Optional | For patients under 1 month of age |

### **Appendix 3** : SwissPedData Common Data Model (CDM), Version 1.0

| Module(s) | Common Data Element | Format | Standardized response options | Importance | Comment / Description |
|---|---|---|---|---|---|
| Infectious diseases | **Maternal syphilis serology** | standardized options | Positive<br>Negative<br>Unknown | Mandatory | For patients under 1 month of age |
| Infectious diseases | **Maternal rubella serology** | standardized options | Positive<br>Negative<br>Unknown | Mandatory | For patients under 1 month of age |
| Infectious diseases | **Maternal toxoplasmosis serology** | standardized options | Positive<br>Negative<br>Unknown | optional | For patients under 1 month of age |
| Infectious diseases | **Maternal Chagas serology** | standardized options | Positive<br>Negative<br>Unknown | Mandatory | For patients under 1 month of age |
| Metabolic diseases | **Self-monitoring of blood glucose** | yes/no | | Optional | Regular self-monitoring of blood glucose done at home |
| Metabolic diseases | **Self-monitoring of ketone bodies** | yes/no | | Optional | Regular self-monitoring of ketone bodies done at home |
| Nephrology | **Prenatal ultrasound** | standardized option | Normal<br>An-/Oligohydramnios<br>Polyhydramnios<br>Megacystis<br>Megaureter<br>Bilateral renal pelvis dilatation > 10 mm<br>Bilateral renal pelvis dilatation < 10 mm<br>Unilateral renal pelvis dilatation > 10 mm<br>Renal cysts<br>Renal agenesis or ectopia<br>Multicystic-dysplastic kidney and bladder extrophy | Mandatory | |
| Nephrology | **Family history of renal disease (1st-2nd degree)** | yes/no | | Mandatory | |
| Neurology | **Seizure type (ILEA 2017 Classification of Seizures)** | standardized options | Focal Onset<br>Generalized Onset<br>Unknown Onset<br>Unclassified | Recommended | Seizure type according to the ILEA 2017 classification of seizures |
| Neurology | **Family history of neurological diseases** | yes/no | | Recommended | Family history of any type of neurological diseases |
| Pulmonology | **Cough** | standardized options | No<br>Yes, acute and dry<br>Yes, acute and wet<br>Yes, chronic and dry<br>Yes, chronic and wet | Recommended | Cut-off for acute/chronic 4 weeks |
| Rheumatology | **Recurrent fever** | yes/no | | Mandatory | History of recurrent fever |

**Appendix 3** : SwissPedData Common Data Model (CDM), Version 1.0

| Module(s) | Common Data Element | Format | Standardized response options | Importance | Comment / Description |
|---|---|---|---|---|---|
| Rheumatology | **History of uveitis** | yes/no | | Mandatory | Presence of active uveitis |
| Rheumatology | **History of inflammatory skin disease** | yes/no | | Mandatory | Presence of skin involvement |
| Rheumatology | **Family history of inflammatory rheumatic disease** | standardized options | No<br>Yes, without spondyloarthropathy<br>Yes, with spondyloarthropathy | Mandatory | Presence of any rheumatic disease in the family |
| Rheumatology | **Family history of inflammatory skin disease** | standardized options | No<br>Yes, without psoariasis<br>Yes, with psoriasis | Mandatory | Presence of any skin disease in the family |
| Rheumatology | **Family history of chronic intestinal diseases** | yes/no | | Mandatory | Presence of any chronic intestinal disease in the family |
| Rheumatology | **Family history of recurrent fever** | yes/no | | Mandatory | Presence of recurrent fever in the family |
| **Domain: Physical examination** | | | | | |
| General paediatrics | **Heart rate** | number | | Mandatory | Heart rate in beats per minute |
| General paediatrics | **Systolic blood pressure** | number | | Mandatory | Value of the systolic blood pressure in mmHg |
| General paediatrics | **Diastolic blood pressure** | number | | Mandatory | Value of the diastolic blood pressure in mmHg |
| General paediatrics | **Respiratory rate** | number | | Mandatory | Respiratory rate in breaths per minute |
| General paediatrics | **Oxygen saturation** | number | | Mandatory | Measured oxygen saturation in % |
| General paediatrics | **Temperature** | number | | Mandatory | Measured temperature of the patient in Celsius degrees |
| General paediatrics | **Weight** | number | | Mandatory | Measured weight of the patient in kg |
| General paediatrics | **Height** | number | | Mandatory | Measured height of the patient in cm |
| General paediatrics | **Head circumference** | number | | Mandatory | Measured head circumference of the patient in cm |
| Endocrinology | **Sitting height** | number | | Recommended | Sitting height measured sitting with straight back in in cm |
| Endocrinology | **Arm span** | number | | Recommended | Arm span: arms streched horizontally, measurement from fingertip to fingertip in cm |
| Endocrinology | **Waist circumference** | number | | Recommended | In cm |
| Endocrinology | **Hip circumference** | number | | Recommended | In cm |
| Endocrinology | **Goiter** | yes/no | | Recommended | Presence of goiter |
| Endocrinology | **Gynecomastia** | standardized options | No<br>Yes, unilateral<br>Yes, bilateral | Recommended | Presence of gynecomastia |
| Endocrinology<br>Metabolic diseases | **Dysmorphic signs** | yes/no | | Recommended | Presence of dysmorphic features. If answer is yes, specification with standardized classification to be defined. |

**Appendix 3** : SwissPedData Common Data Model (CDM), Version 1.0

| Module(s) | Common Data Element | Format | Standardized response options | Importance | Comment / Description |
|---|---|---|---|---|---|
| Endocrinology | **Cryptorchidism** | standardized options | No<br>Yes, unilateral<br>Yes, bilateral | Mandatory | Presence of cryptorchidism |
| Endocrinology | **Insulin injection site** | standardized options | Normal<br>Abnormal, lipoatrophy<br>Abnormal, lipohypertrophy | Optional | Inspection of insulin delivery sites |
| Endocrinology | **Retinopathy screening** | normal/abnormal | | Optional | |
| Endocrinology | **Neuropathy screening performed** | standardized options | No<br>Yes, vibration<br>Yes, monofilament | Optional | |
| Endocrinology | **Testis volume right side** | number | | Mandatory | Volume of right testis in ml |
| Endocrinology | **Testis volume left side** | number | | Mandatory | Volume of left testis in ml |
| Endocrinology | **Tanner breast stage** | number | | Mandatory | |
| Endocrinology | **Tanner pubic hair stage** | number | | Mandatory | |
| Endocrinology | **Tanner axillary hair stage** | number | | Mandatory | |
| Endocrinology | **Tanner genital stage** | number | | Recommended | |
| Endocrinology | **Breast size** | number | | Optional | In cm |
| Endocrinology | **Female genital examination** | normal/abnormal | | Mandatory | |
| Endocrinology | **Penis length** | number | | Recommended | In cm |
| Endocrinology | **Chovstek sign** | yes/no | | Recommended | Twitching of facial muscles in response to tapping over the area of the facial nerve |
| Endocrinology | **Trousseau sign** | yes/no | | Recommended | Carpopedal spasm that results from ischemia |
| Endocrinology | **Thyroid nodule** | yes/no | | Recommended | Presence of thyroid nodule |
| Infectious diseases<br>Metabolic diseases<br>Rheumatology | **Hepatomegaly noted at physical examination** | yes/no | | Mandatory | |
| Infectious diseases<br>Metabolic diseases<br>Rheumatology | **Splenomegaly noted at physical examination** | yes/no | | Mandatory | |
| Infectious diseases | **Meningeal signs noted at physical examination** | yes/no | | Mandatory | |
| Infectious diseases<br>Rheumatology | **Skin lesion noted at physical examination** | yes/no | | Mandatory | |
| Infectious diseases | **Irritability noted during physical examination** | yes/no | | Mandatory | |
| Infectious diseases<br>Rheumatology | **Adenopathy noted at physical examination** | standardized options | No<br>Yes, localized | Mandatory | |

**Appendix 3** : SwissPedData Common Data Model (CDM), Version 1.0

| Module(s) | Common Data Element | Format | Standardized response options | Importance | Comment / Description |
|---|---|---|---|---|---|
| | | | Yes, generalized | | |
| Infectious diseases | **Respiratory distress noted at physical examination** | yes/no | | Mandatory | |
| Infectious diseases | **Conjunctivitis noted at physical examination** | yes/no | | Mandatory | |
| Infectious diseases | **Prolonged capillary refill time (> 2 sec) noted at physical examination** | yes/no | | Mandatory | |
| Infectious diseases | **Signs of dehydration noted at physical examination** | standardized options | No<br>Yes, < 5%<br>Yes, 5-10%<br>Yes, >10% | Mandatory | |
| Metabolic diseases | **Skin abormalities** | yes/no | | Mandatory | Presence of skin abnormalities |
| Metabolic diseases | **Abnormal body proportions** | yes/no | | Recommended | Presence of abnormal body proportions |
| Nephrology | **Average 24-hour arterial pressure, systolic** | number | | Mandatory | In mmHg |
| Nephrology | **Average 24-hour arterial pressure, diastolic** | number | | Mandatory | In mmHg |
| Nephrology | **Average daytime systolic BP** | number | | Mandatory | In mmHg |
| Nephrology | **Average daytime diastolic BP** | number | | Mandatory | In mmHg |
| Nephrology | **Average night-time systolic BP** | number | | Mandatory | In mmHg |
| Nephrology | **Average night-time diastolic BP** | number | | Mandatory | In mmHg |
| Nephrology | **Mean Arterial Pressure (MAP)** | number | | Mandatory | Measured MAP in mmHg |
| Nephrology | **Blood pressure dipping pattern** | number | | Mandatory | Difference between daytime mean systolic pressure and night-time mean systolic pressure expressed as a percentage of the day value |
| Neurology | **Walking ability** | standardized options | Community ambulator<br>Household ambulator<br>Non-ambulatory | Mandatory | |
| Pulmonology | **Auscultation** | normal/abnormal | | Mandatory | |
| Pulmonology | **Thorax shape** | normal/abnormal | | Mandatory | The shape of the thorax |
| Rheumatology | **Active arthritis** | yes/no | | Mandatory | Presence of active arthritis |
| Rheumatology | **If active arthritis: number of joints involved** | number | | Mandatory | Number of joints involved in active arthritis |
| Rheumatology | **Maximal mouth opening** | number | | Mandatory | Maximal mouth opening in mm |
| Rheumatology | **Muscle strength** | normal/abnormal | | Recommended | Overall muscle strength |

**Appendix 3** : SwissPedData Common Data Model (CDM), Version 1.0

| Module(s) | Common Data Element | Format | Standardized response options | Importance | Comment / Description |
|---|---|---|---|---|---|
| **Domain: Clinical scores** | | | | | |
| General paediatrics | **Triage scale (ED), type** | standardized options | Australasian Triage Scale<br>Canadian Triage Scale<br>Other | Mandatory | Name of the triage scale used |
| General paediatrics | **Triage scale (ED), value** | number | | Mandatory | Value of the triage scale |
| General paediatrics | **AVPU score** | standardized options | Alert<br>Voice<br>Pain<br>Unresponsive | Mandatory | |
| General paediatrics | **Glasgow Coma Scale** | number | | Mandatory | |
| Cardiology | **Modified Ross heart failure classification for children** | standardized options | Class I<br>Class II<br>Class III<br>Class IV | Mandatory | Class I: Asymptomatic<br>Class II: Mild tachypnea or diaphoresis with feeding in infants, dyspnea on exertion in older children<br>Class III: Marked tachypnea or diaphoresis with feeding in infants, marked dyspnea on exertion, prolonged feeding times with growth failure<br>Class IV: Symptoms such as tachypnea, retractions, grunting or diaphoresis at rest |
| Cardiology | **NYHA classification for adults** | standardized options | Class I<br>Class II<br>Class III<br>Class IV | Mandatory | Class I: No symptoms and no limitation in ordinary physical activity<br>Class II: Mild symptoms (mild shortness of breath and/or angina) and slight limitation during ordinary activity.<br>Class III: Marked limitation in activity due to symptoms, even during less-than-ordinary activity. Comfortable only at rest.<br>Class IV: Severe limitations. Experiences symptoms even while at rest. Mostly bedbound patients. |
| Endocrinology | **Endocrinology clinical score type** | standardized options | Crook score<br>Billewicz score<br>Ferriman-Gallway score<br>Prader stage<br>External genitalia score | Optional | Type of score |
| Endocrinology | **Endocrinology clinical score result** | number | number | Optional | Result of score |
| Gastroenterology | **PCDAI** | number | | Mandatory | Paediatric Crohn's Disease Activity Index |
| Gastroenterology | **PUCAI** | number | | Mandatory | Paediatric Ulcerative Colitis Activity Index |
| Gastroenterology | **PYMS score** | number | | Mandatory | Paediatric Yorkhill Malnutrition Score |
| Gastroenterology | **Bristol stool scale** | number | | Mandatory | |
| Allergy/Immunology | **SCORAD index** | number | | Mandatory | SCORing Atopic Dermatitis Index |
| Metabolic diseases<br>Neurology | **Developmental test: Type** | standardized options | Bayley II<br>Bayley III | Mandatory | Type of developmental test performed |

| Module(s) | Common Data Element | Format | Standardized response options | Importance | Comment / Description |
|---|---|---|---|---|---|
| | | | Griffith<br>Other | | |
| Metabolic diseases<br>Neurology | **Development test: Results** | normal/abnormal | | Mandatory | Result of developmental test performed |
| Metabolic diseases | **Developmental delay** | yes/no | | Mandatory | Developmental delay as assessed by treating physician |
| Nephrology | **CKD stage** | number | | Mandatory | Chronic Kidney Disease stage |
| Pulmonology | **Epworth Sleepiness Scale** | number | | Mandatory | |
| Pulmonology | **Lung-to-Head-Ratio** | number | | Mandatory | Congenital diaphragmatic hernia |
| Pulmonology | **PICADAR** | number | | Mandatory | PrImary CiliARy DyskinesiA Rule |
| **Domain: Investigations** | | | | | |
| General paediatrics | **Type of radiological study (detailed)** | standardized options | See comments | Mandatory | Standard classification to be defined |
| General paediatrics | **Date and time of imaging study** | datetime | YYYY-MM-DD hh:mm:ss | Mandatory | Date and time of the radiological study |
| General paediatrics | **Radiation dose** | number | | Mandatory | If applicable, dose of radiation in mSv |
| General paediatrics | **Indication for the imaging study** | free text | | Mandatory | Medical reason for the radiological study |
| Cardiology | **ECG performed** | yes/no | | Mandatory | Date of study should be recorded |
| Cardiology | **Holter-ECG** | yes/no | | Mandatory | Date of study should be recorded |
| Cardiology | **Ergometry** | yes/no | | Mandatory | Date of study should be recorded |
| Cardiology | **Echocardiography performed** | yes/no | | Mandatory | Detailed standardized echo measurements will be discussed in the future. Date of study should be recorded |
| Cardiology | **Cardiac electrophysiology study performed** | yes/no | | Mandatory | Date of study should be recorded |
| Cardiology | **Diagnostic cardiac catheterization (hemodynamic study) performed** | yes/no | | Mandatory | Date of study should be recorded |
| Endocrinology | **Bone age: method** | standardized options | Greulich & Pyle<br>BoneXpertR<br>Tanner Whitehouse | Mandatory | Method used to assess radiographic bone age. Date of study should be recorded |
| Endocrinology | **Bone age: result** | number | | Mandatory | Bone age result in years |
| Endocrinology<br>Metabolic diseases | **Use of continuous glucose monitoring** | yes/no | | Recommended | Use of glucose sensor |
| Endocrinology | **Number of days per week with continuous glucose monitoring** | number | | Mandatory | Days per week |

**Appendix 3** : SwissPedData Common Data Model (CDM), Version 1.0

| Module(s) | Common Data Element | Format | Standardized response options | Importance | Comment / Description |
|---|---|---|---|---|---|
| Endocrinology | **Continuous glucose monitoring: Device** | standardized options | Freestyle libre<br>Freestyle libre 2<br>Dexcom G5<br>Dexcom G6<br>Medtronic Guardian<br>Medtronic Enlyte | Mandatory | |
| Endocrinology | **Blood glucose self-measurement** | number | | Mandatory | Number of measures per week |
| Endocrinology | **Scans per day** | number | | Mandatory | If Flash Glucose Monitoring (FGM) is used |
| Endocrinology | **Blood ketone measurement** | number | | Mandatory | Number of measures per week |
| Endocrinology | **Mean glucose** | number | | Mandatory | mmol/l |
| Endocrinology | **Glucose variability** | number | | Mandatory | % |
| Endocrinology | **Time in range** | number | | Mandatory | Time between 4.0 and 10.0 mmol/l in % |
| Endocrinology | **Time in hypoglycemia** | number | | Mandatory | Time < 3.9mmol/l in % |
| Gastroenterology | **Type of gastrointestinal endoscopy** | standardized options | Upper<br>Lower<br>Upper and lower<br>Other | Mandatory | Date of study should be recorded |
| Gastroenterology | **Indication for gastrointestinal endoscopy** | standardized options | Rectal bleeding<br>Abdominal pain<br>Dysphagia<br>Diarrhea<br>Other | Mandatory | Medical reason for the endoscopic study. Other include for example oesophageal atresia or other anatomical abnormality, food impaction |
| Gastroenterology | **Gastrointestinal endoscopic biopsy** | yes/no | | Mandatory | Gastrointestinal endoscopic biopsy performed. Date of study should be recorded |
| Gastroenterology | **Impedance-pHmetry** | yes/no | | Mandatory | Date of study should be recorded |
| Gastroenterology | **Type of breath test** | standardized options | Lactose<br>Lactulose<br>Fructose<br>Urea<br>Other | Mandatory | Type of breath test. Date of study should be recorded |
| Gastroenterology | **Capsule endoscopy** | yes/no | | Mandatory | Date of study should be recorded |
| Gastroenterology | **Endoscopic ultrasound** | yes/no | | Mandatory | Date of study should be recorded |
| Gastroenterology | **Liver biopsy** | yes/no | | Mandatory | Date of study should be recorded |
| Allergy/Immunology | **Prick-test performed** | yes/no | | Mandatory | Date of study should be recorded |
| Allergy/Immunology<br>Pulmonology | **sIgE performed** | yes/no | | Mandatory | sIgE stands for specific serum immunoglobulin E. Date of study should be recorded |
| Allergy/Immunology | **Result of sIgE** | positive/negative | | Mandatory | sIgE stands for specific serum immunoglobulin E |

**Appendix 3** : SwissPedData Common Data Model (CDM), Version 1.0

| Module(s) | Common Data Element | Format | Standardized response options | Importance | Comment / Description |
|---|---|---|---|---|---|
| Allergy/Immunology | **Result of prick-test** | positive/negative | | Mandatory | Date of study should be recorded |
| Allergy/Immunology | **Allergen challenge performed** | yes/no | | Mandatory | Date of study should be recorded |
| Allergy/Immunology | **Allergen challenge result** | positive/negative | | Mandatory | |
| Infectious diseases | **Urine collection method** | standardized options | Urethral catheterization<br>Clean catch void<br>Urine collection bag<br>Mid-stream urine<br>Suprapubic aspiration | Mandatory | Method of collection of urine for culture. Date of study should be recorded |
| Infectious diseases | **Mantoux test** | number | | Mandatory | In mm. Date of study should be recorded |
| Infectious diseases | **Mantoux test: interpretation** | standardized options | Positive<br>Negative<br>Doubtful<br>Unknown | Mandatory | Healthcare provider’s interpretation of Mantoux test |
| Infectious diseases | **IGRA result** | standardized options | Positive<br>Negative<br>Indeterminate | Mandatory | IGRA stands for Interferon-Gamma Release Assay. Date of study should be recorded |
| Nephrology | **Renal ultrasound result** | normal/abnormal | | Mandatory | Date of study should be recorded |
| Nephrology | **Renal MRI result** | normal/abnormal | | Mandatory | Date of study should be recorded |
| Nephrology | **Voiding cystourethrography or kidney microbubble ultrasound results** | standardized options | No vesicoureteral reflux<br>Vesicoureteral reflux, unilateral – Grade I<br>Vesicoureteral reflux, unilateral – Grade II<br>Vesicoureteral reflux, unilateral – Grade III<br>Vesicoureteral reflux, unilateral – Grade IV<br>Vesicoureteral reflux, bilateral – Grade I<br>Vesicoureteral reflux, bilateral – Grade II<br>Vesicoureteral reflux, bilateral – Grade III<br>Vesicoureteral reflux, bilateral – Grade IV | Mandatory | Date of study should be recorded |
| Nephrology | **Posterior urethral valves** | yes/no | | | |
| Nephrology | **Renal scintigraphy results** | standardized options | Normal<br>Hypoplasia<br>Scars<br>Other | Mandatory | Date of study should be recorded |
| Nephrology | **Estimated GFR by Schwartz formula** | number | | Mandatory | GFR [ml/min] |

**Appendix 3** : SwissPedData Common Data Model (CDM), Version 1.0

| Module(s) | Common Data Element | Format | Standardized response options | Importance | Comment / Description |
|---|---|---|---|---|---|
| Nephrology | **Proteinuria** | number | | Mandatory | In mg/mmol (spot-urine) or mg/m2/h for 24h Urine |
| Nephrology<br>Neurology<br>Pulmonology | **Genetic test performed** | yes/no | | Mandatory<br>Recommended | Mandatory for nephrology, recommended for neurology and pulmonology |
| Neurology | **Neurologic electrophysiologic study: Type** | standardized options | EEG<br>EMG<br>AEP<br>SEP<br>VEP<br>Other | Mandatory | EEG: electroencephalogram, EMG: electromyography, AEP: auditory evoked potentials, SEP: somatosensory evoked potentials, VEP: visual evoked potential<br>Date of study should be recorded |
| Neurology | **Neurologic electrophysiologic study: Result** | normal/abnormal | | Mandatory | |
| Metabolic diseases<br>Neurology | **Hearing test: Type** | standardized options | OAE<br>AEP<br>Pure tone audiometry | Mandatory | OAE: otoacoustic emissions, AEP: auditory evoked potentials<br>Date of study should be recorded |
| Metabolic diseases<br>Neurology | **Hearing test: Result** | normal/abnormal | | Mandatory | |
| Neurology | **Vision test: Performed by** | standardized options | Ophtalmologist<br>Optometrist<br>Peadiatrician<br>Other | Recommended | Health professional who tested vision |
| Neurology | **Vision test: Result** | normal/abnormal | | Recommended | |
| Neurology | **Lumbar puncture performed** | yes/no | | Mandatory | Date of study should be recorded |
| Neurology | **Opening Pressure at Lumbar Puncture** | number | | Optional | Opening pressure in cmH2O |
| Pulmonology | **Spirometry performed** | yes/no | | Mandatory | Date of study should be recorded |
| Pulmonology | **Lung function: RV** | number | | Recommended | RV: Residual volume. In L |
| Pulmonology | **DLCO** | number | Diffusion capacity of the lung for carbon monoxide | Recommended | DLCO: diffusing capacity of the lungs for carbon monoxide. In ml CO/min/mmHg |
| Pulmonology | **Lung function: Bronchodilator administered** | yes/no | | Mandatory | |
| Pulmonology | **Bronchoscopy performed** | yes/no | | Mandatory | Date of study should be recorded |
| Pulmonology | **Lung function: Challenge test performed (treadmill, methacholine challenge test)** | yes/no | | Mandatory | |
| Pulmonology | **Broncho-alveolar lavage performed** | yes/no | | Mandatory | |

**Appendix 3** : SwissPedData Common Data Model (CDM), Version 1.0

| Module(s) | Common Data Element | Format | Standardized response options | Importance | Comment / Description |
|---|---|---|---|---|---|
| Pulmonology | **Sweat test results** | standardized options | | Mandatory | Chloride in mmol/l (Macroduct)<br>Conductivity in mmol/l eq NaCl (Nanoduct)<br>Date of study should be recorded |
| Pulmonology | **Sleep studies** | standardized options | Polysomnography<br>Respiratory Polygraphy<br>Oximetry | Mandatory | Sleep studies performed<br>Date of study should be recorded |
| Pulmonology | **Lung function: FEV1** | number | | Mandatory | FEV1: forced expiratory volume-one second. pre/post absolute number in L |
| Pulmonology | **Lung function: FVC** | number | | Mandatory | FVC: forced vital capacity. Pre/post. In L |
| Pulmonology | **Lung function: TLC** | number | | Recommended | TLC: Total lung capacity. In L |
| Pulmonology | **Lung function: LCI** | number | | Recommended | LCI: Lung clearance index. Equipment / gas currently in use in each center |
| Pulmonology | **Lung function: Nasal NO** | number | | Recommended | Nasal NO: Nasal nitric oxide measurement. in ppb or nl/mn |
| Pulmonology | **Lung function: FeNO** | number | | Recommended | FeNO: exhaled nitric oxide test. Online or Off-line method. absolute number in ppb |
| Pulmonology | **Lung function: CPET performed** | yes/no | | Recommended | CPET: Cardiopulmonary Exercise Testing |
| Pulmonology | **Lung function: FEF 25-75** | number | | Recommended | FEF25-75: Forced expiratory flow over the middle one half of the FVC (force vital capacity). in L/s |
| Pulmonology | **Lung function: FEV 0.75** | number | | Recommended | FEV 0.75: forced expiratory volume in 3/4 of a second. pre/post. absolute number in L |
| Pulmonology | **Lung function: sRaw** | number | | Recommended | sRaw: specific airway resistance. kPa/sec |
| Pulmonology | **Lung function: FRC** | number | | Recommended | FRC: functional residual capacity. in L |
| Pulmonology | **Lung function: FRC: Test** | standardized options | Bodyplethysmography<br>MBW | Recommended | MBW: multiple breath washout |
| **Domain: Diagnosis** | | | | | |
| General paediatrics | **Diagnosis** | See comments | See comments | Mandatory | Inpatients diagnosis are ICD10 coded and outpatients diagnosis are free text. |
| General paediatrics | **Date of diagnosis** | date | YYYY-MM-DD | Mandatory | |
| General paediatrics | **Cause of death** | See comments | See comments | Mandatory | Standard classification to be defined |
| General paediatrics | **Date of death** | date | YYYY-MM-DD | Mandatory | |
| Cardiology | **IPCCC diagnosis** | standardized options | IPCCC Code | Mandatory | IPCCC: International Paediatric and Congenital Cardiac Code |
| Allergy/Immunology | **Allergic disease confirmation** | standardized options | Skin prick test<br>Allergen challenge<br>sIgE | Mandatory | |
| Infectious diseases | **If infectious diagnosis: Type of documentation** | standardized options | Clinically documented infection<br>Microbiologically documented infection | Mandatory | |

**Appendix 3** : SwissPedData Common Data Model (CDM), Version 1.0

| Module(s) | Common Data Element | Format | Standardized response options | Importance | Comment / Description |
|---|---|---|---|---|---|
| Infectious diseases | **If infectious diagnosis: Nosocomial** | yes/no | | Mandatory | |
| Infectious diseases | **If nosocomial infection: Date of first symptom** | date | YYYY-MM-DD | Mandatory | |
| Infectious diseases | **If nosocomial infection: Site of infection** | standardized options | Respiratory tract<br>Gastro-intestinal tract<br>Urinary tract<br>Surgical site<br>Other | Mandatory | |
| Metabolic diseases | **Diagnosis confirmation** | standardized options | Clinical<br>Biochemical<br>Enzymatic<br>Genetic | Mandatory | The way diagnosis has been confirmed |
| Metabolic diseases | **Diagnosis suspicion** | standardized options | Prenatal<br>Newborn<br>Selective | Mandatory | The type of screening that led to the diagnosis |
| Neurology | **OMIM code** | standardized options | OMIM code | Recommended | OMIM: Online Mendelian Inheritance in Man |
| Neurology | **HPO code** | standardized options | HPO code | Optional | HPO: Human Phenotype Ontology |
| **Domain: Treatment** | | | | | |
| General paediatrics | **Drug name** | standardized options | International non-proprietary name | Mandatory | Name of the drug(s) received as inpatient |
| General paediatrics | **Prescribed drug at discharge** | standardized options | International non-proprietary name | Mandatory | Name of the drug(s) prescribed at discharge |
| General paediatrics | **Route of administration** | standardized options | Oral<br>Intravenous<br>Subcutaneous<br>Intramuscular<br>Intrathecal<br>Rectal<br>Inhalation<br>Cutaneous<br>Ocular<br>Nasal<br>Otic<br>Other | Mandatory | |
| General paediatrics | **Date and time of first administration** | datetime | YYYY-MM-DD hh:mm:ss | Mandatory | Time of first administration of the drug |
| General paediatrics | **Date and time of last administration** | datetime | YYYY-MM-DD hh:mm:ss | Mandatory | Time of last administration of the drug |
| General paediatrics | **Frequency of administration** | number | | Mandatory | Number of administrations per 24 hours |
| General paediatrics | **Dose** | number | | Mandatory | Dose given at each administration of the drug |

**Appendix 3** : SwissPedData Common Data Model (CDM), Version 1.0

| Module(s) | Common Data Element | Format | Standardized response options | Importance | Comment / Description |
|---|---|---|---|---|---|
| General paediatrics | **Dose unit** | standardized options | | Mandatory | List of possible units to be defined |
| General paediatrics | **Reason for discontinuation of treatment** | standardized options | Recovery<br>Change to another medication<br>No effect observable<br>Adverse events<br>Reducing polypharmacy<br>Other | Mandatory | Reason why a treatment is stopped |
| General paediatrics | **Adverse events** | standardized options | MedDRA classification | Mandatory | MedDRA: Medical Dictionary for Regulatory Activities |
| General paediatrics | **Supplemental O2: Date and time of start** | datetime | YYYY-MM-DD hh:mm:ss | Mandatory | Time at starting oxygen therapy |
| General paediatrics | **Supplemental O2: Date and time of interruption** | datetime | YYYY-MM-DD hh:mm:ss | Mandatory | Time at stopping oxygen therapy |
| General paediatrics | **Supportive services: Type** | standardized options | Physiotherapy<br>Ergotherapy<br>Social assistance<br>Other | Mandatory | |
| Endocrinology | **Type of insuline therapy** | standardized options | MDI<br>CSII | Mandatory | MDI: Multiple dose injection. CSII: Continuous subcutaneous insulin infusion |
| Endocrinology | **Total daily dose of insuline (long and short acting)** | number | | Mandatory | units per kg per day |
| Endocrinology | **Basal insuline** | number | | Mandatory | Percentage of basal insuline (%) |
| Gastroenterology<br>Metabolic diseases | **Therapeutic diet** | yes/no | | Mandatory | Therapeutic diet prescribed by physician |
| Metabolic diseases | **Type of therapeutic diet** | standardized options | Low-protein<br>Ketogenic<br>Low-fat<br>Frequent meals<br>Nocturnal feed<br>Medical food<br>Other | Mandatory | Type of therapeutic diet prescribed |
| Allergy/Immunology | **Epinephrine Pen prescribed** | yes/no | | | |
| Infectious diseases | **BCG immunization** | standardized options | Yes<br>No<br>Unknown | Mandatory | |
| Neurology | **Rehabilitation supportive devices: Type** | standardized options | Upper limb orthoses<br>Lower limb orthoses<br>Corset<br>Standing frame<br>Walking aid (crutches NF-walker, rollator etc.)<br>Wheelchair: Manual | Recommended | |

**Appendix 3** : SwissPedData Common Data Model (CDM), Version 1.0

| Module(s) | Common Data Element | Format | Standardized response options | Importance | Comment / Description |
|---|---|---|---|---|---|
| | | | Wheelchair: Electric powered<br>Other | | |
| Pulmonology | **Pulmonary rehabilitation** | yes/no | | Recommended | |
| **Domain: Equipment and procedures** | | | | | |
| General paediatrics | **Equipment type** | standardized options | See comments | Mandatory | Standard classification to be defined |
| General paediatrics | **Equipment date of insertion** | date | YYYY-MM-DD | Mandatory | |
| General paediatrics | **Equipment date of withdrawal** | date | YYYY-MM-DD | Mandatory | |
| Cardiology | **Cardiac procedures** | standardized options | IPCCC Code | Mandatory | IPCCC: International Paediatric and Congenital Cardiac Code |
| Cardiology | **Date of cardiac procedure** | date | YYYY-MM-DD | Mandatory | Date of intervention |
| Gastroenterology | **Therapeutic gastrointestinal endoscopic procedures** | standardized options | Haemostasis<br>Oesophageal dilatation (Balloon/Savary)<br>Percutaneous endoscopic gastrostomy (PEG)<br>Endoscopic retrograde cholangiopancreatography (ERCP)<br>Other | Mandatory | |
| Nephrology | **Type of dialysis (1)** | standardized options | Acute<br>Chronic | Mandatory | |
| Nephrology | **Type of dialysis (2)** | standardized options | Haemodialysis<br>Peritoneal dialysis<br>Hemodiafiltration | Mandatory | If peritoneal dialysis, type of catheter and number of peritonitis should be specified |
| Nephrology | **Date of dialysis initiation** | datetime | YYYY-MM-DD hh:mm:ss | Mandatory | |
| Nephrology | **Date of dialysis termination** | datetime | YYYY-MM-DD hh:mm:ss | Mandatory | |
| Nephrology | **Dialysis: vascular access type** | standardized option | Central venous catheter<br>Arteriovenous fistula<br>Arteriovenous graft | Mandatory | If central venous catheter, its localization should be specified |
| Nephrology | **Renal transplantation, graft (1)** | standardized options | Deceased donor<br>Living donor | Mandatory | |
| Nephrology | **Renal transplantation, graft (2)** | standardized options | Related donor<br>Unrelated donor | Mandatory | |
| Nephrology | **Renal transplantation** | standardized options | Preemptive transplantation<br>Nonpreemptive transplantation | Mandatory | |
| Nephrology | **Renal transplantation: Number of received grafts** | number | | Mandatory | Number of grafts received including present one |
| Nephrology | **Plasmapheresis performed** | yes/no | | Mandatory | |

| Module(s) | Common Data Element | Format | Standardized response options | Importance | Comment / Description |
|---|---|---|---|---|---|
| Nephrology | **Renal biopsy performed** | standardized options | No<br>Yes, without complication in the following 24 hours<br>Yes, with complications in the following 24 hours | Mandatory | |
| Nephrology | **Cystoscopy performed** | yes/no | | Mandatory | |
| Nephrology | **Angiography performed** | yes/no | | Mandatory | |
